# A Genes & Health pilot recall study of intrahepatic cholestasis of pregnancy and cholestatic liver disease

**DOI:** 10.1101/2024.10.18.24314654

**Authors:** Maria Constantinides, Joseph Gafton, Ana Cristina Angel Garcia, Genes and Health, Peter H. Dixon, Catherine Williamson, Kenneth Linton, Sarah Finer, Upkar S. Gill, Julia Zöllner

## Abstract

**Background:** Cholestatic liver disease disproportionately affects South Asians, yet they remain underrepresented in genomic studies. This recall study aimed to recall volunteers from a British South Asian genetic cohort that were considered to be at high risk of cholestatic liver disease based on their genotype or phenotype.

**Methods:** Cases were defined as participants with rare (minor allele frequency <1%) heterozygous loss of function (LoF) variants in *ABCB4* and *ABCB11* (genotype re-call) or with a previous intrahepatic cholestasis of pregnancy (ICP) diagnosis (ICD10 O26.6). Cases were matched 1:1 to controls. A detailed medical and family history was taken along with fasting anthropometric and transient elastography (TE) measurements and blood samples.

**Results:** Out of 22 eligible volunteers, 9 (41%) participated in the recall (8/9 genotype and 1/9 phenotype recall). Among the cases there were 5 *ABCB4* LoF, 3 *ABCB11* LoF, and 1 ICP phenotype. Of these, 6/9 (66.7%) were newly identified with a cholestatic phenotype (genotype re-call). Specifically, 3/6 (50%) had increased liver stiffness on TE with one also demonstrating abnormal liver blood tests. 2/6 (33.3%) experienced at least 2 cholestatic symptoms and an additional 1/6 (16.7%) demonstrated abnormal liver blood tests without increased liver stiffness.

**Conclusion:** This pilot study demonstrated new evidence of cholestatic liver disease in 66.7% of volunteers, underscoring the potential of rare heterozygous *ABCB4/11* variants as markers for identifying individuals at high risk of developing cholestatic liver disease. Consequently, individuals at higher genetic risk benefit from monitoring, personalised treatment and prevention strategies for cholestatic liver disease.

**Plain language summary:** We aimed to identify British South Asians at high risk of liver disease due to specific genetic factors, such as issues with bile production or liver problems during pregnancy. We invited these individuals to a clinic, where we collected their medical and family history, conducted liver blood tests, and performed a scan to check for early signs of liver scarring. We found that nearly two out of three participants had undetected liver disease. This finding suggests that genetic factors are linked to developing liver disease, highlighting the importance of early detection and monitoring for those at risk.

## Introduction

Cholestatic liver disease is a growing cause of morbidity and mortality worldwide as it can lead to liver fibrosis and liver cirrhosis (1–3). In the United Kingdom, there has been a 400% increase in mortality due to liver disease over the last 50 years with an acceleration in liver disease rates compared with other major diseases (4). In South Asian populations liver diseases are more common, although they often remain undiagnosed and under-investigated (5,6). Cholestatic liver disease encompasses a broad range of diseases characterised by jaundice and cholestasis which can result in end-stage liver disease, cirrhosis, and other-severe liver-related complications. Symptoms of cholestatic liver disease include pruritus, abdominal pain (epigastric or right upper quadrant pain (RUQ)), steatorrhea, jaundice, dark urine and pale stools (1). Cholestasis biochemically is characterised by an increase in alkaline phosphatase (ALP) or gammalZIglutamyl transferase (GGT) an increase in bilirubin, which can occur at a later stage due to decrease in bile flow and, when measured, elevated serum bile acid concentrations (7).

Genes involved in bile formation such as *ABCB4* (encodes canalicular phosphatidylcholine floppase) and *ABCB11 (*encodes bile salt export pump (BSEP)) have been associated with cholestatic liver disease (8)(9)(10). Homozygous variants in *ABCB4* and *ABCB11* are associated with a variety of phenotypes ranging from mild cholestasis to severe familial cholestasis such as progressive familial intrahepatic cholestasis (PFIC)(11). Specifically, *ABCB4* variants have been shown to predispose to adult biliary cirrhosis, gallstone, gallbladder and bile duct carcinoma, drug induced cholestasis and low phospholipid associated cholelithiasis (LPAC) (10)(11)(12)(13)(14). It appears that carriers of heterozygous variants in genes associated with disease can develop cholestatic disease with a spectrum in the disease phenotype and onset (11,15). For example, in genetically susceptible individuals the altered physiological state of pregnancy, that is accompanied by an increase in serum concentrations of oestrogen and progesterone and their metabolites, can reveal a cholestatic phenotype (i.e. intrahepatic cholestasis of pregnancy (ICP), the commonest gestational liver disease) (16). The cause of ICP is multifactorial but a study showed that 12.8% of mothers, 15.9% of sisters and 10.3% of daughters were affected, highlighting a strong genetic link (17). Furthermore, a European ancestry study demonstrated that 20% of severe ICP cases harboured a pathogenic/likely pathogenic heterozygous variant in *ABCB4* or *ABCB11*(18). Not only is ICP associated with adverse pregnancy outcomes (11), women with ICP also have an increased risk of developing hepatobiliary diseases such liver cirrhosis, hepatitis C, cholangitis and gallstones (18–23). In actual fact, the presence of *ABCB4* variants in ICP patients increased the risk of developing hepatobiliary disease in an Icelandic population (24). Similarly, our recent work identified novel variants associated with cholestatic liver disease and demonstrated an increased heterozygous rare variant burden associated with ICP in a British Bangladeshi and Pakistani population (11). For the treatment of cholestasis, ursodeoxycholic acid (UDCA) is the most commonly used drug with other medications such as rifampicin, cholestyramine and fibrates being used (25). Medical management with UDCA is considered to be the first line therapy in all cases with PFIC, with some cases with PFIC benefiting from surgical biliary diversion or ileal bile acid inhibitor treatment (26,27), with liver transplantation being used when treatment fails (14)(25)(28).

Genes & Health is a community-based genetic study of health and disease in British Bangladeshi and Pakistani people, who are underrepresented in existing cohorts. This study recalls existing volunteers already recruited to Genes & Health and in whom linkage to health and genetic data is already established. The individuals recalled were previously identified by our group (11) to be at high risk of cholestatic liver disease based on their genotype (rare heterozygous loss of function (LoF) variants in *ABCB4* or *ABCB11*) or phenotype (ICP). Our aim was to identify cases at highest genetic risk of cholestatic liver disease and correlate genetic risk with clinical findings. We hypothesised that heterozygous LoF variants or a previous history of ICP would predispose to cholestatic liver disease and may be detectable in advance of symptomatic disease onset.

## Methods

### Genes & Health Study

Genes & Health volunteers are currently recruited in three sites across the United Kingdom (East London, Bradford and Manchester), in community (mosques, markets and libraries) and health care (NHS GP practices, outpatient clinics) settings. DNA (saliva) is taken for SNP (single nucleotide polymorphism) array genotyping and exome sequencing (29). Genes & Health combines electronic health record data, with primary and secondary care records and genetic data as previously described (29).

### Study Design

A recall-by-genotype and a recall-by-phenotype approach was implemented in Genes & Health participants with exome sequencing data available. The study was conducted between April 2023 – August 2023. At the time of the study, exome sequencing data was available for 5236 volunteers reporting parental relatedness. Participants were recruited and seen in East London Genes & Health.

### Ethical Approval

Genes & Health ethical approval was granted by the South East London National Research Ethics Committee (14/LO/1240) in 2014. This recall study operated with ethical approval from the West of Scotland Research Ethics Service (22/WS/0109).

### Participant Recruitment

Figure 1 describes targeted recruitment to this recall study. Cases with rare (minor allele frequency (MAF) < 1%) heterozygous LoF variants in genes highly associated with disease - *ABCB4* and *ABCB11* (see supplementary Table 1) were recalled. LoF variants impair the functionality of human protein-encoding genes. Homozygous LoF variants are associated with severe cholestatic liver disease phenotypes, therefore heterozygous mutations with ensuing haploinsufficiency are likely to increase susceptibility to disease and were considered high risk for the development of a sub-clinical cholestatic liver disease phenotype. Additional cases were selected as those with ICP phenotype (coded through ICD10 O26.6 in electronic health records) with either rare heterozygous single nucleotide variants (SNV) or LoF variants (see supplementary Table 2).

**Figure 1.**
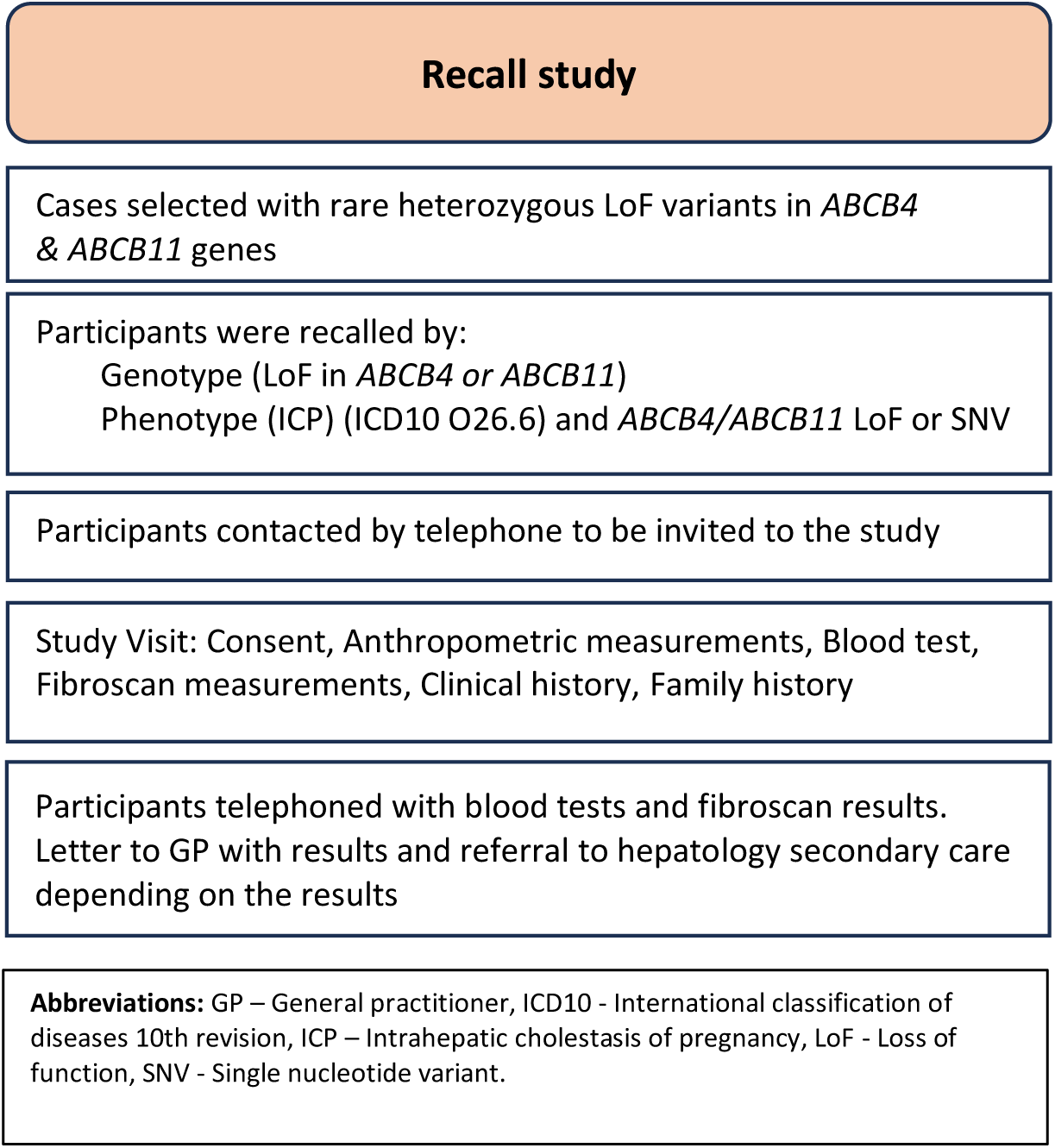
Re-call study participant selection.

Cases were matched with a 1:1 ratio to healthy controls. Controls were Genes & Health volunteers matched according to age (+/- 5 years), sex, and ethnicity (British Pakistani or British Bangladeshi). Exclusion criteria for controls were anyone with pathogenic variant in *ABCB4/ABCB11/ATP8B1* (genes associated with cholestatic liver disease), known cholestatic liver disease, liver cirrhosis or carcinoma, bile duct carcinoma, hepatitis B/hepatitis C, gallstones, covid infection in the preceding 6 weeks, any new medication introduced in the last 3 months causing cholestatic side effects such as jaundice, dark urine, pale stools, pruritus.

### Study Visit Appointment

Participants fasted for a minimum of 8 hours. A detailed medical history and family history was taken by a clinician (MC or JG). At the visit anthropometric measurements (height, weight, waist circumference and hip circumference) and bioimpedance were taken. Blood analysis included plasma aliquots and blood cell RNA preservation (Paxgene). Quantitative analysis included full blood count, urea and electrolytes, bone profile, liver blood tests (alanine transaminase (ALT), aspartate transaminase (AST), GGT, enhanced liver fibrosis (ELF) test (30), bile acid concentration, virology screen (Hepatitis B surface Ag (HBsAg), Hepatitis C IgG, HIV 1/2 antibodies), lipids, fasting glucose and glycated haemoglobin (Hba1c). Routine biochemical blood results were incorporated into Fibrosis 4 index (FIB-4) score (31). For FIB-4, a score of 1.3 was used to rule out advanced fibrosis and a score of 3.25 was used to rule in advanced fibrosis (32).

To assess hepatic fibrosis transient elastography (TE) using FibroScan compact 530 (Echosens) was performed. Values of liver stiffness measurement (LSM, in kPa)((surrogate marker of liver fibrosis) (33) and liver CAP (in dB/m) (surrogate marker of liver steatosis) (34) were obtained. When measuring LSM to ensure reliable results IQR/Median (%) shall remain ≤ 30% (35). No quality criteria have been defined when measuring CAP. As per EASL guidelines(32), LSM < 8 kPa was used to rule out fibrosis, LSM 10 – 12 kPa indicating probable advanced chronic liver disease and LSM > 12-15 kPa to rule in compensated advanced chronic liver (cACLD). Cut off points differ with aetiology but provide an overall approximation (32). Although there has not been a general consensus for cut-off values, CAP values above 275 dB/m were used to indicate steatosis as per EASL guidelines (32). We classified participants as having new evidence of cholestatic liver disease if they had abnormal TE with or without cholestatic symptoms or abnormal liver blood tests, experiencing two or more cholestatic symptoms or abnormal liver blood tests alone. Cholestatic symptoms were defined as pruritus, RUQ/epigastric pain, jaundice, dark urine, pale stools and steatorrhea.

### Statistical Analysis

Statistical analysis was performed using SPSS for Mac Version 28 and 29 and statistical significance was set at p<0.05. Normality of data was assessed using histograms and the Shapiro Wilk test. Continuous variables following normal distribution were analysed using independent t-test. Continuous variables not following normal distribution were analysed using the Mann-Whitney Test. Nominal data were analysed using Fischer’s exact test.

### Data Availability

Genetic summary data of the participants including exome sequencing results are available on the genes & health website (https://www.genesandhealth.org/research/scientific-data-downloads).

## Results

Out of 22 eligible cases, nine (41%) attended a clinic appointment (6 females, and 3 male cases) that were matched to 9 controls (Table 1). Among the cases there were 5 *ABCB4* LoF, 3 *ABCB11* LoF, and 1 ICP phenotype. Figure 2 illustrates the cases who attended the clinic for recall and their variants. The clinical and demographic characteristics of cases and controls are shown in Table 2. No statistically significant difference was found in the demographics between cases and controls.

**Figure 2.**
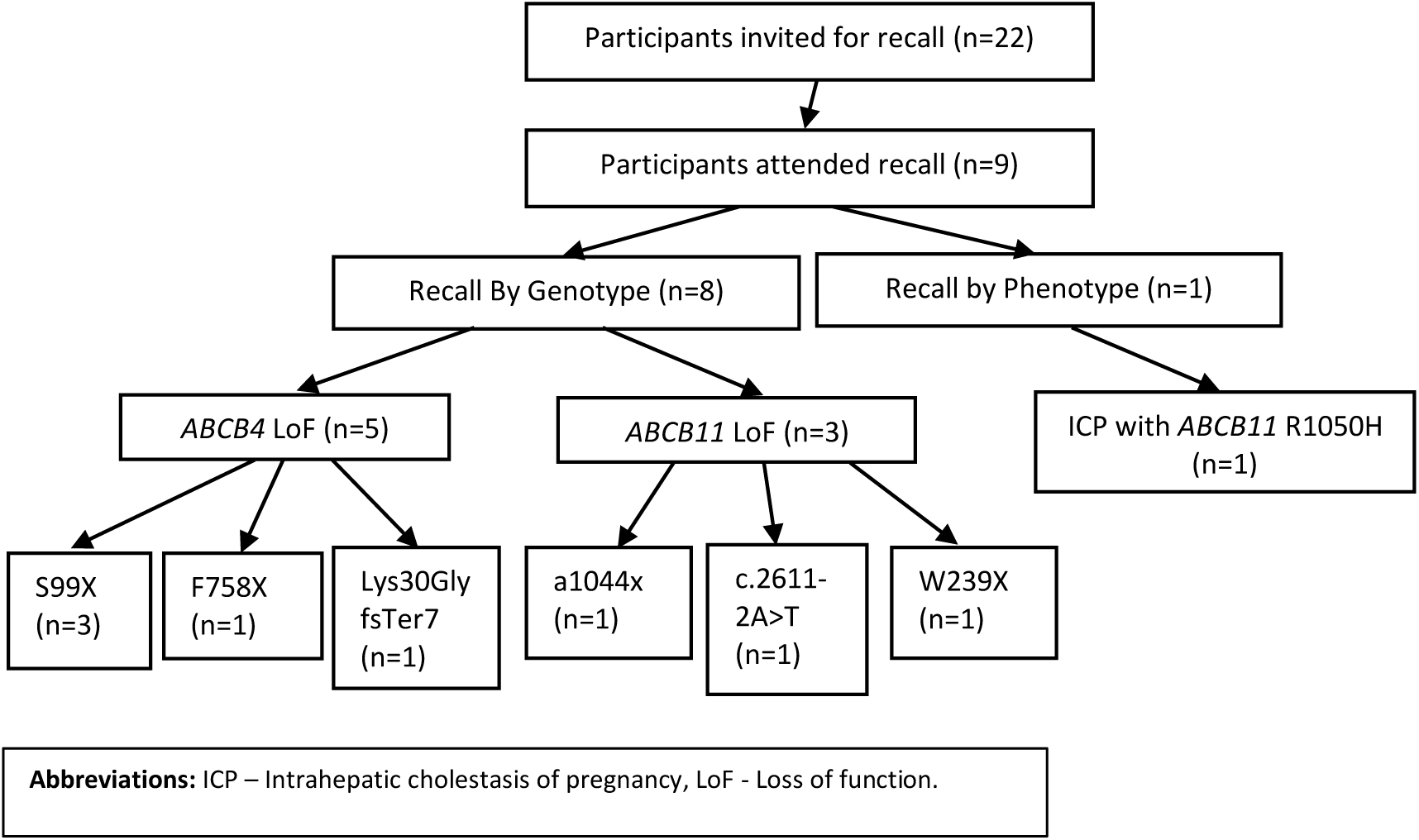
Schematic flow chart of participants that attended recall (n=number of volunteers)

**Table 1.** Genotype and phenotype used to select cases for recall.

| Recall by genotype |  |  |  |  |  |  |  |  |
| --- | --- | --- | --- | --- | --- | --- | --- | --- |
| Volunteers (n) | Gene | Type | Variants | Effect | Zygosity | Transcript | GnomAD AF | G&H AF |
| 3 | *ABCB4* | LoF | S99x | Frameshift | Het | ENSP00000395716.1: p. Ser99LeufsTer11 | 0.00039970 | 0.00336538 |
| 1 | *ABCB4* | LoF | F758X | Frameshift | Het | ENSP00000392983.1: p. Leu759TyrfsTer38 | . | 0.00009610 |
| 1 | *ABCB4* | LoF | Lys30GlyfsTer7 | Frameshift | Het | ENSP00000392983.1: p. Lys30GlyfsTer7 | 0.00000408 | 0.00020243 |
| 1 | *ABCB11* | LoF | A1044x | Frameshift | Het | ENSP00000497931.1: p. Ala1044LeufsTer53 | . | 0.00009562 |
| 1 | *ABCB11* | LoF | c.2611-2A>T | Splice-acceptor variant | Het | ENST00000263817.7: c.2611- 2A>T | 0.00000407 | 0.00057870 |
| 1 | *ABCB11* | LoF | W239x | Stop gained | Het | ENSP00000497931.1: p. Trp239Ter | . | 0.00009566 |
| Recall by phenotype (ICP) |  |  |  |  |  |  |  |  |
| 1 | *ABCB11* | SNV | R1050H | - | Het | ENSP00000497931.1: p. Arg1050His | 0.00000421 | 0.00019135 |

**Abbreviations:** Het – Heterozygous, ICP – intrahepatic cholestasis of pregnancy, LoF - Loss of function, SNV – Single nucleotide variant.

**Table 2.** Baseline characteristics of re-call participants.

| Characteristic | Cases (n=9) | Controls (n=9) | p value |
| --- | --- | --- | --- |
| Age |  |  |  |
| years | 41.00 (14.5) | 44.00(14.5) | 0.792 |
| min, max | 34, 57 | 32, 53 | - |
| BMI (kg/m<sup>2</sup>) | 31.31(10.21) | 28.70 (6.78) | 0.310 |
| Waist-Hip Ratio | 0.90 (0.05) | 0.90 (0.13) | 0.734 |
| Fat percentage (%) | 34.90 (12.55) | 33.0 (15.85) | 0.350 |
| Gender |  |  |  |
| Female | 6 (66.7%) | 6 (66.7%) | 1.000 |
| Ethnicity |  |  |  |
| British Pakistani | 6 (66.7%) | 6 (66.7%) | 1.000 |
| British Bangladeshi | 3 (33.3%) | 3 (33.3%) | 1.000 |
| Parity |  |  |  |
| Parous | 5 (83.3%) | 6(100%) | 1.000 |

**Abbreviations:** BMI – Body mass index.
Age, BMI, Waist-Hip Ratio, Fat percentage are presented as Median (IQR, Interquartile range) and p-value obtained using Mann Whitney test. Gender, ethnicity, parity are presented as number (%) and p value was obtained using Fisher's exact test. A p-value <0.05 was considered statically significant.

### Existing Diagnoses

Table 3 describes the clinical and biochemical findings in cases and controls. Out of the 9 cases, 4/9 (44.4%) had a known diagnosis of metabolic dysfunction associated steatotic liver disease (MASLD) of whom 3/4 (75%) had an *ABCB4* LoF and 1/4 (25%) had an *ABCB11* LoF variant (Table 4). 2/9 (22.2%) had a known diagnosis of gallstones both of whom had an *ABCB11* variant (Table 4). No participant with *ABCB4* LoF variants was found to have gallstones.

**Table 3.** Clinical characteristics of cases and controls.

| Investigations | Cases (n=9) | Controls (n=9) | P –Value |
| --- | --- | --- | --- |
| **Blood tests** |  |  |  |
| ALP | 77.0 (41.50) | 74.0 (39.50) | 0.399 |
| ALT | 30.0 (24.50) | 24.0 (11.0) | 0.111 |
| AST | 22.0 (13.0) | 20.0 (10.0) | 0.594 |
| **GGT** | **26.0 (36.0)** | **19.0 (8.0)** | **<0.01\*** |
| Bilirubin | 6.0 (6.0) | 6.0 (3.0) | 0.964 |
| AFP | 1.80 (3.05) | 1.40 (1.65) | 0.097 |
| LDH | 146 (31.5) | 201 (59.0) | 0.058 |
| Bile acids | 0.1 (3.90) | 0.1 (3.40) | 0.783 |
| ELF score | 8.57 (1.09) | 8.79 (1.67) | 0.860 |
| **Serum markers of fibrosis** |  |  |  |
| FIB-4 | 0.80 (0.196) | 0.875 (0.20) | 0.457 |
| **Transient elastography** |  |  |  |
| **CAP (dB/m) – Mean** | **297.00 (67.37)** | **238.25 (33.66)** | **0.021\*** |
| LSM (kPA) – Mean | 6.68 (4.25) | 4.28 (1.11) | 0.072 |
| Evidence of Severe Fibrosis (F3) | 3/9 | 0/8 # | 0.124 |
| Evidence of Steatosis (>275 dB/m) | 6/9 | 2/8 # | 0.109 |

**Abbreviations:** AFP - Alpha-fetoprotein, ALP - Alkaline phosphatase, ALT - Alanine transaminase, AST - Aspartate transaminase, CAP - Controlled attenuation parameter, , ELF – Enhanced liver fibrosis, FIB-4 – Fibrosis-4 score, GGT – Gamma-glutamyl transferase, LDH - Lactate dehydrogenase, LSM – Liver stiffness measurement.
Data are presented as Median (IQR) or Mean (SD). P value obtained either using t test, Mann Whitney test or Fischer’s exact test. \*A p-value <0.05 was considered statically significant.

**Table 4.** Clinical and biochemical findings in cases recalled by genotype and phenotype.

|  | LoF | Demographics | BMI | Background | ALT (IU/L) | ALP (IU/L) | GGT (IU/L) | BA (umol/l) | TE - LSM (kPa) (IQR/med) | TE - CAP (dB/m) | Cholestatic Symptoms (Y/N) | Maternal Disease (Y/N) | F/H of Biliary Disease (Y/N) | Evidence of sub-clinical cholestatic disease (Y/N) |
| --- | --- | --- | --- | --- | --- | --- | --- | --- | --- | --- | --- | --- | --- | --- |
| **A**<br>**B**<br>**C**<br>**B**<br>**4** | S99X | 36-40F G3P1 | 31.5 | NAFLD<br>Pre-diabetes<br>IDA | 33 | 54 | 26 | 6 | 4.6 (22%) | **357 (17)** | Y – RUQ pain | Y - PET | N | N |
|  | S99X | 46-50F G0P0 | 36.1 | NAFLD<br>PCOS<br>Hypercholesterolemia<br>Fibroids<br>Primary infertility | 33 | 74 | 27 | <2 | **11.6 (38%)** | **292 (34)** | N | N | Y- Gallstones (Parent's sibling) | Y – Abnormal TE |
|  | S99X | 36-40F G3P2 | 39.4 | No past medical history | 20 | 73 | 30 | <2 | 4.8 (6%) | 253 (32) | N | N | Y- Gallstones (Parent's sibling) | N |
|  | Lys30GlyfsTer7 | 51-55M | 31.3 | T2DM<br>HTN | **60** | 107 | **91** | 8 | **10.2 (25%)** | **372 (34)** | Y – Itching, Dark Urine, RUQ pain | N/A | N | Y – Abnormal LFTs, Abnormal TE |
|  | F758x | 31-35M | 29.6 | NAFLD<br>Renal calculus | **210** | **159** | **267** | 2 | 4.3 (16%) | **334 (48)** | Y - Epigastric pain | N/A | N | Y – Abnormal LFTs |
| **A**<br>**B**<br>**C**<br>**B**<br>**1**<br>**1** | a1044x | 56-60F G9P7 | 31.0 | NAFLD<br>Gallstones<br>Cholecystectomy<br>HTN<br>Hypercholesterolaemia<br>T2DM | 30 | 82 | 24 | <2 | 4.8 (23%) | 188 | Y - Itching in all pregnancies (x7), Steatorrhea, RUQ and epigastric pain | Y - Itching during all pregnancies | Y - Itching during pregnancy (x3 children), died of 'liver problems' (parent) | Y – Cholestatic symptoms |
|  | c.2611-2A>T | 41-45M | 23.3 | HTN<br>Hypercholesterolaemia<br>Renal calculus | 24 | 119 | 25 | <2 | 3.0 (17%) | 220 | Y – RUQ, and Epigastric pain<br>Steatorrhea | N/A | N | Y – Cholestatic symptoms |
|  | W239x | 46-50F G4P4 | 42.1 | HTN<br>T2DM | 17 | 70 | 26 | 2 | **14.3 (26%)** | **373** | Y - Itching during 2nd pregnancy, RUQ/ and epigastric pain | Y - Itching during 2nd pregnancy<br>GDM | Y- Gallstones (Parent) | Y – Abnormal TE, Cholestatic symptoms |
| **I** | ABC B11 SNV | 36-40F G2P2 | 25.5 | Gallstones<br>ICP | 30 | 77 | 21 | <2 | 2.4 (8%) | **284** | Y – Acute episode of | Y – ICP<br>PET | Y – Acute episode of | Y – Cholestatic |
| C<br>P<br>P<br>H<br>E<br>N<br>O<br>T<br>Y<br>P<br>E | R10<br>50H |  |  |  |  |  |  |  |  |  | jaundice,<br>itching<br>during<br>pregnancy<br>(ICP) |  | cholestasi<br>s and liver<br>abnormali<br>ty during<br>pregnancy<br>(Parent),<br>died of<br>'hepatitis'<br>(Parent's<br>Siblings –<br>x3) | symptoms<br>,<br>ICP<br>diagnosis |

**Abbreviations:** ALT - Alanine transaminase (normal range – 10-50 IU/L ), ALP - Alkaline phosphatase (normal range – 40-129 IU/L ), BA – Bile acids (normal range - <10 umol/l, in pregnancy <19-20 umol/l non-fasting), BMI – Body mass index , CAP - Controlled attenuation parameter , F/H – Family history, F- Female, GGT - Gamma glutamyl transferase (normal range 10-71 IU/L), HTN – Hypertension, IDA – Iron deficiency anaemia, LFT – liver function test, LoF – Loss of function, M – Male, NAFLD – Non-alcoholic fatty liver disease, PCOS – Polycystic ovary syndrome, PET – Preeclampsia, RUQ – right upper quadrant, SNV, single nucleotide variant, T2DM – Type 2 diabetes mellitus, TE – Transient elastography.

### Blood Tests

Overall, a statistically significant difference was found between cases and controls in the level of GGT. No statistically significant difference was found in other biochemical markers of cholestatic liver disease or clinical fibrosis scores (Table 3). Figure 3 illustrates the distribution of ALP, AST, ALT and GGT between participants with *ABCB4* and *ABCB11* variants. 2/9 (22%) cases had elevated liver transaminases _[WC1]_ compared to none of the controls. One of the two cases with raised concentrations of liver transaminases, also had elevated ALP levels (Table 4). Both cases with high liver transaminase concentrations had an *ABCB4* LoF variant. No cases with *ABCB11* LoF variants had abnormal liver blood tests.

**Figure 3.**
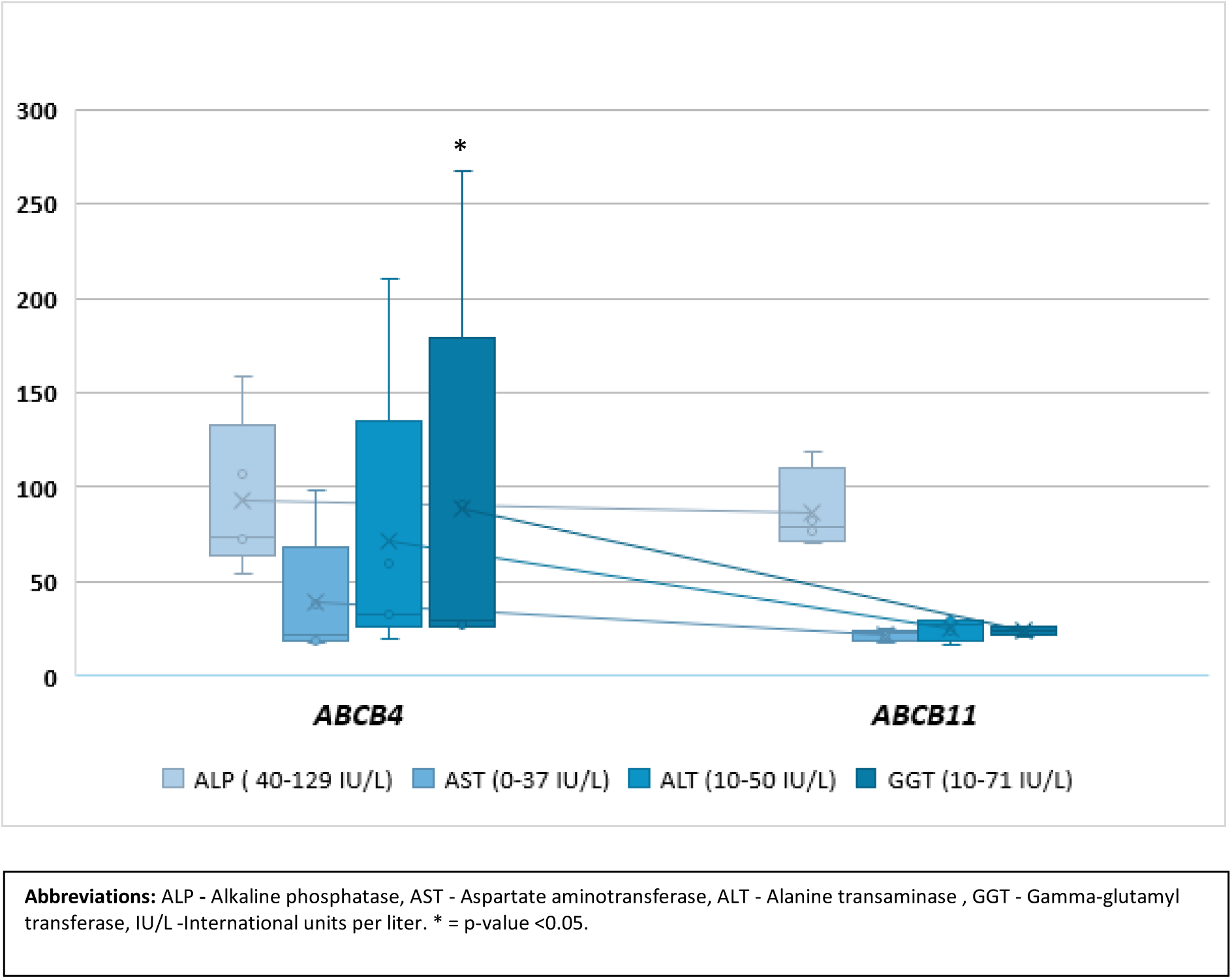
Box Whisker plot illustrating liver blood tests in participants with *ABCB4* and *ABCB11 variants*.

### Cholestatic Symptom*s*

Overall, 7/9 (77.7%) cases either reported active or had a previous history of cholestatic symptoms (Table 4). There were 6 cases who experienced cholestatic symptoms at the time of the recall, 3/6 (50%) cases experienced one cholestatic symptom and 3/6 (50%) experienced two or more cholestatic symptoms. The most common cholestatic symptom reported was RUQ pain or epigastric pain (6/6) followed by steatorrhea (2/6), itching (1/6), and dark urine (1/6) (Table 4).

Of the 5 parous females, 1/5 (20%) had a confirmed diagnosis of ICP (recalled based on phenotype and the presence of an *ABCB11* SNV R1050H variant). In her first pregnancy, she experienced severe itching with bile acid concentrations exceeding 400 µmol/L, necessitating treatment with UDCA. She underwent an emergency caesarean section at 33 weeks, and the neonate required a 21-day stay in the neonatal intensive care unit (NICU). In her second pregnancy, she was started early on UDCA, with bile acid concentration reaching about 400 µmol/L. This time, she delivered via elective caesarean section at 36 weeks with no NICU admission. Additionally, this participant had a jaundice episode after starting a progesterone only pill. Further, 2/5 (40%) participants had a history suggestive of ICP, reporting significant itching during pregnancy despite no formal diagnoses. In contrast, none of the participants in the control group had cholestatic symptoms during pregnancy or at the time of recall.

### Evidence of Cholestatic Liver Disease

Hepatic fibrosis was identified in 3/9 (33.3%) cases but not in any controls (Table 4). 2/3 (66.6%) of cases with hepatic fibrosis had an *ABCB4* LoF and 1/3 (33.3%) had an *ABCB11* LoF variant (Table 4).

Hepatic steatosis was identified in 6/9 (66.7%) cases and 2/8 (25%) controls. 4/6 (66.6%) of cases with hepatic steatosis had an *ABCB4* LoF) and 2/6 (33.3%) had an *ABCB11* LoF or SNV (Table 4). A statistically significant difference between cases and controls was found in the mean CAP (p values = 0.021), indicating a higher degree of steatosis in cases (Table 3).

Overall, 6/9 (66.7%) cases were classified with new evidence of cholestatic disease. 3/6 (50%) were classified with new evidence of cholestatic disease due to abnormal liver stiffness measurements (LSM) on transient elastography (Table 3 and 4). They presented with LSM values between 10 – 14.3kPa indicating probable or confirmed compensated advanced chronic liver (cACLD).

The remaining 2/6 (33.3%) were classified due to currently experiencing two or more cholestatic symptoms (Table 4). 1/6 (16.7%) was classified due to abnormal liver blood tests) (Table 4). No control was found to have evidence of cholestatic liver disease. Figure 4 illustrates amongst the cases the relationship between genotype, cholestatic symptoms, blood tests, and fibroscan results.

**Figure 4.**
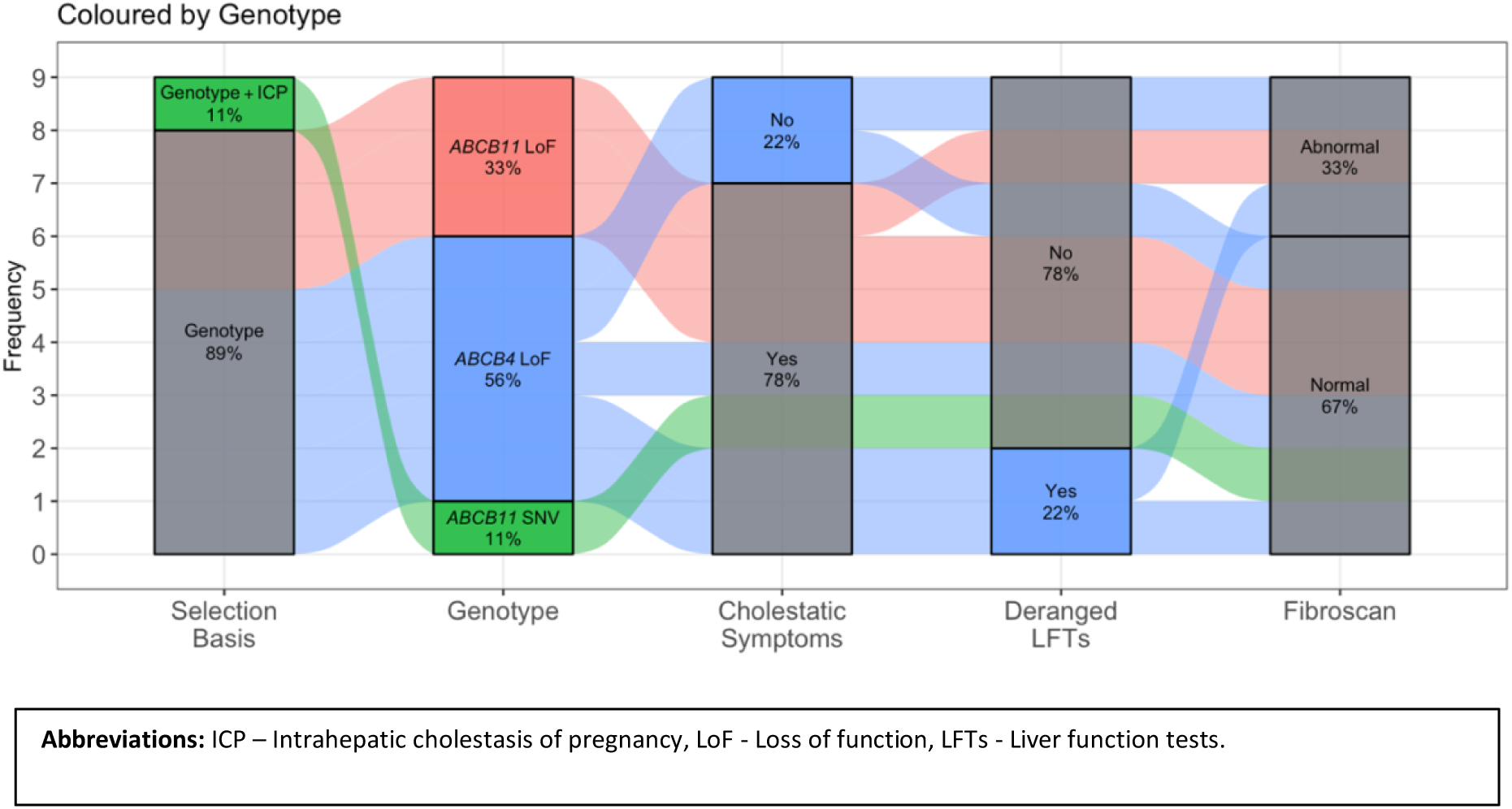
Graph illustrating the different outcomes according to genotype recall or phenotype recall.

## Discussion

In this study we report the results of the first gene candidate recall by genotype and phenotype study associated with cholestatic liver disease in a unique cohort of British Bangladeshi and British Pakistani subjects. Owing to the unique structure of Genes & Health, we were able to recall participants with rare genetic variants suspected to be at high risk of cholestatic liver disease and assess their phenotype. We were able to contribute to the limited evidence base that exists on how heterozygous variants in *ABCB4* and *ABCB11* manifest phenotypically (11)(15)(36). We hypothesised that high risk genetic variants predispose to cholestatic liver disease. We were able to demonstrate that over half of cases (66.7%) had new evidence of cholestatic liver disease either due to evidence of fibrosis on TE scan, abnormal liver tests or cholestatic symptoms. With the decreasing cost of genotyping, there arises a promising opportunity to conduct targeted genotyping for individuals at elevated risk of future cholestatic liver diseases, such as patients with ICP and their relatives. We are optimistic that our findings will serve as a catalyst for further genotype-based recall studies. Such studies aim to identify asymptomatic individuals showing signs of cholestatic disease early, enabling timely interventions that may prevent disease progression. Prophylactic administration of UDCA may decelerate fibrosis progression and reduce the risk of subsequent complications in some affected individuals (28). With the growth of various biobanks, there is an emerging opportunity to implement prophylactic UDCA treatment for individuals identified as high-risk due to *ABCB4* or *ABCB11* variants. This proactive approach could potentially mitigate the development of cholestatic liver disease. There have also been reports of an in vitro study using targeted pharmacotherapy with ivacaftor (used in the treatment of cystic fibrosis) rescuing the function of some variants of *ABCB11* (37). It has also been shown that prophylactic administration of UDCA in some of the affected individuals may slow down fibrosis progression and delay a risk of potential complications (28). For people with variants in hepatobiliary transporters, e.g. *ABCB4*, detected following an ICP diagnosis Hagenbeck et al. recommend lifelong use of UDCA, annual ultrasound studies and monitoring laboratory parameters with the aim of preventing long term sequalae (20)(38).

### Phenotype

The application of TE in the general population without known liver disease remains limited, although its utility in a tertiary setting is well-established. According to EASL, non-invasive tests like the FIB-4 index are recommended initially for patients at risk of chronic liver disease in primary care settings. Those with a FIB-4 score of ≥1.30 should undergo liver stiffness measurement using TE. In our study, despite no participant having a FIB-4 score above 1.30, we identified three cases with abnormal liver stiffness measurements. As per EASL guidelines genetic testing is recommended after exclusion of more frequent causes of cholestatic liver disease in adults (39). This context is particularly relevant given our findings that 44.4% of the individuals we recalled were previously diagnosed with MASLD. Interestingly, Nayagam et al. who identified four patients with *ABCB4* variants and MASLD, observed that the two patients who underwent a biopsy exhibited biliary disease without fatty liver disease histologically. Consequently, the original MASLD diagnosis was reconsidered. This finding prompts us to question whether our participants had an accurate diagnosis of MASLD, especially since none underwent a liver biopsy to confirm their conditions.

Although in the literature there is a reported association between *ABCB4* variants and LPAC, none of our cases with *ABCB4* LoF were found to have gallstones (13)(40). Whereas previous studies reported no association between *ABCB11* pathogenic variants and gallstones (21), we report two cases of *ABCB11* (LoF a 1044x, SNV R1050H) with gallstones. Our findings add to the existing body of research on *ABCB11* variants and gallstones, as previously documented in seven Dutch patients. These patients, who presented with gallstones and benign recurrent intrahepatic cholestasis type 2, carried missense and splice site variants of the gene(41).

The sole participant formally diagnosed with ICP was recalled due to their phenotype and an *ABCB11* SNV (R1050H). Despite experiencing two severe episodes of ICP, an episode of jaundice after taking a progesterone-only pill, and having gallstones, there was no evidence of cholestatic liver disease present at the time of recall. Monrose et al. reported that the median time from an ICP diagnosis to the onset of liver disease is approximately 13.1 years, underscoring the importance of dedicated long-term follow-up for these patients (42). Although the participant in our study currently showed no signs of liver disease, they remain at elevated risk for developing cholestatic liver disease in the future. In our cohort, while no other parous females were formally diagnosed with ICP, two of the five parous females—both harbouring *ABCB11* LoF variants—exhibited symptoms suggestive of ICP. Interestingly, none of the parous females with *ABCB4* LoF mutations presented with similar symptoms. This observation contrasts with existing literature that suggests *ABCB4* variants have a greater overall genetic influence on ICP susceptibility than *ABCB11* variants (10). However, it is important to note that most studies to date have focused on populations of European descent. Consequently, our findings may reflect ancestry-specific variations in disease etiology, highlighting the need for further research in diverse populations.

### Cholestatic Symptoms and Blood Tests

Our findings reveal distinct clinical features associated with *ABCB4* or *ABCB11* variants among our participants. Notably, we observed that two participants with *ABCB4* variants had abnormal liver blood tests, including elevated levels of ALT, AST, GGT and ALP. In contrast, all participants carrying *ABCB11* variants displayed cholestatic symptoms with no evidence of abnormal liver blood tests. Additionally, a large-scale whole-genome sequencing of the Icelandic population showed an association between *ABCB4* rare variants and increased levels of AST, ALT and GGT, further supporting our observations (24). Abnormal liver blood tests in our study can be potentially explained by *ABCB4* haploinsufficiency, which impedes the neutralisation of bile salts due to reduced biliary phospholipids, thereby damaging the canalicular membrane (25)(43). This is consistent with what has been reported in literature that *ABCB4* variants are characterised by higher levels of GGT compared to *ABCB11* (25). However, as illustrated by other studies, GGT levels alone may not reliably differentiate between cholestasis linked to *ABCB4* and *ABCB11* variants, since some *ABCB4* variants may not show elevated GGT levels (25). Our observations align with recent research indicating that heterozygous *ABCB4* variants are frequently seen in adults with cholestasis. Notably, a study in Switzerland found that *ABCB4* variants were present in 50% of individuals assessed for unexplained biochemical cholestasis, ICP, or other cholestatic phenotypes. These findings underline the prevalent role of *ABCB4* in various forms of cholestasis and highlight the importance of considering genetic backgrounds when diagnosing and managing these conditions (36).

### Limitations

This is a small study focussing on genes known to play a role in the aetiology of cholestatic liver disease (11). We adopted a pragmatic approach to recall *ABCB4* and *ABCB11* LoF variants, given their established impact on protein function. While this approach was tailored to our study’s objectives, it may have introduced some selection bias. We were only able to recall one participant with previous history of ICP. As reported in other recall by genotype (RbG) studies, the small sample size of participants we recalled can lead to a higher variance in results compared with a larger cohort (44). However, we were able to achieve a 41% recall in a disadvantaged and high risk group which makes this study stand out. Compared to other recall studies, we included a matched control group to allow a direct comparison and reduce the risk of variance. To standardise our approach, we asked participants to be fasted. However, this can affect bile acids as mean, non-fasting serum bile acid concentrations are higher than in fasted individuals (45). In addition, most non-invasive tests such as serum markers of hepatic fibrosis and transient elastography were developed and validated in secondary or tertiary settings and have not been tested for use in primary care or the general population (32). To evaluate steatosis, TE was used to determine CAP however ultrasound remains the first line for steatosis detection (33).

## Conclusion

In summary, we report the first recall-by genotype and phenotype study of individuals identified to be at high risk of cholestatic liver disease based on their genotype or phenotype. The majority of cases (66.7%) had new evidence of liver disease or symptoms of cholestatic liver disease. Notably, we demonstrated how by first identifying individuals to be at high risk of cholestatic disease by genotype, exploring their phenotype and performing investigations such as liver blood tests and TE scans we were able to demonstrate evidence of cholestatic liver disease and arrange appropriate investigations and follow up. As a result, we hope that integration of genetic information can potentially facilitate personalised medicine according to genotype to make the best therapeutic choice for each individual.

## Acknowledgments

Genes & Health is/has recently been core-funded by Wellcome (WT102627, WT210561), the Medical Research Council (UK) (M009017, MR/X009777/1, MR/X009920/1), Higher Education Funding Council for England Catalyst, Barts Charity (845/1796), Health Data Research UK (for London substantive site), and research delivery support from the NHS National Institute for Health Research Clinical Research Network (North Thames). Genes & Health is/has recently been funded by Alnylam Pharmaceuticals, Genomics PLC; and a Life Sciences Industry Consortium of AstraZeneca PLC, Bristol-Myers Squibb Company, GlaxoSmithKline Research and Development Limited, Maze Therapeutics Inc, Merck Sharp & Dohme LLC, Novo Nordisk A/S, Pfizer Inc, Takeda Development Centre Americas Inc.

We thank Social Action for Health, Centre of The Cell, members of our Community Advisory Group, and staff who have recruited and collected data from volunteers. We thank the NIHR National Biosample Centre (UK Biocentre), the Social Genetic & Developmental Psychiatry Centre (King’s College London), Wellcome Sanger Institute, and Broad Institute for sample processing, genotyping, sequencing and variant annotation.

This work uses data provided by patients and collected by the NHS as part of their care and support. This research utilised Queen Mary University of London’s Apocrita HPC facility, supported by QMUL Research-IT, http://doi.org/10.5281/zenodo.438045

We thank: Barts Health NHS Trust, NHS Clinical Commissioning Groups (City and Hackney, Waltham Forest, Tower Hamlets, Newham, Redbridge, Havering, Barking and Dagenham), East London NHS Foundation Trust, Bradford Teaching Hospitals NHS Foundation Trust, Public Health England (especially David Wyllie), Discovery Data Service/Endeavour Health Charitable Trust (especially David Stables), Voror Health Technologies Ltd (especially Sophie Don), NHS England (for what was NHS Digital) - for GDPR-compliant data sharing backed by individual written informed consent.

Most of all we thank all of the volunteers participating in Genes & Health.

A favourable ethical opinion for the main Genes & Health research study was granted by NRES Committee London - South East (reference 14/LO/1240) on 16 Sept 2014. Queen Mary University of London is the Sponsor.

## Notes

### Competing Interest Statement

SF receives research funding for Genes & Health from MRC, NIHR, Alnylam Pharmaceuticals, Takeda, Glaxo Smith Kline, Merck, Pfzer, NovoNordisk, Maze Pharmaceuticals, Bristol Myers Squibb.CW consults for Mirum and GSK.

### Funding Statement

This study did not receive any funding.

## References

1. Onofrio FQ, Hirschfield GM. The Pathophysiology of Cholestasis and Its Relevance to Clinical Practice. Clin Liver Dis. 2020;15(3):110–4.

2. Lu L. Guidelines for the Management of Cholestatic Liver Diseases (2021). J Clin Transl Hepatol [Internet]. 2022;10(4):757–69. Available from: https://www.doi.org/10.14218/JCTH.2022.00147

3. Pollock G, Minuk GY. Diagnostic considerations for cholestatic liver disease. J Gastroenterol Hepatol. 2017;32(7):1303–9.

4. Williams R, Aspinall R, Bellis M, Camps-Walsh G, Cramp M, Dhawan A, et al. Addressing liver disease in the UK: a blueprint for attaining excellence in health care and reducing premature mortality from lifestyle issues of excess consumption of alcohol, obesity, and viral hepatitis. Lancet [Internet]. 2014 Nov;384(9958):1953–97. Available from: https://linkinghub.elsevier.com/retrieve/pii/S0140673614618389

5. Alazawi W, Mathur R, Abeysekera K, Hull S, Boomla K, Robson J, et al. Ethnicity and the diagnosis gap in liver disease: A population-based study. Br J Gen Pract. 2014;64(628):e694–702.

6. Szanto KB, Li J, Cordero P, Oben JA. Ethnic differences and heterogeneity in genetic and metabolic makeup contributing to nonalcoholic fatty liver disease. Diabetes, Metab Syndr Obes. 2019;12:357–67.

7. Mawardi M, Alalwan A, Fallatah H, Abaalkhail F, Hasosah M, Shagrani M, et al. Cholestatic liver disease: Practice guidelines from the Saudi Association for the Study of Liver diseases and Transplantation. Saudi J Gastroenterol. 2021;27(7):S1–26.

8. Wang DQ-H, Portincasa P, Wang HH. Bile Formation and Pathophysiology of Gallstones. In: Encyclopedia of Gastroenterology [Internet]. Elsevier; 2020. p. 287–306. Available from: https://linkinghub.elsevier.com/retrieve/pii/B9780128012383658610

9. Nayagam JS, Foskett P, Strautnieks S, Agarwal K, Miquel R, Joshi D, et al. Clinical phenotype of adult-onset liver disease in patients with variants in ABCB4, ABCB11, and ATP8B1. Hepatol Commun. 2022;6(10):2654–64.

10. Dixon PH, Sambrotta M, Chambers J, Taylor-Harris P, Syngelaki A, Nicolaides K, et al. An expanded role for heterozygous mutations of ABCB4, ABCB11, ATP8B1, ABCC2 and TJP2 in intrahepatic cholestasis of pregnancy. Sci Rep [Internet]. 2017;7(1):1–8. Available from: 10.1038/s41598-017-11626-x

11. Zöllner J, Finer S, Linton KJ, Akhtar S, Anwar M, Arciero E, et al. Rare variant contribution to cholestatic liver disease in a South Asian population in the United Kingdom. Sci Rep. 2023;13(1):1–14.

12. Gonzales E. Liver diseases related to MDR3 (ABCB4) gene deficiency. Front Biosci [Internet]. 2009;Volume(14):4242. Available from: https://imrpress.com/journal/FBL/14/11/10.2741/3526

13. Rosmorduc O, Poupon R. Low phospholipid associated cholelithiasis: Association with mutation in the MDR3/ABCB4 gene. Orphanet J Rare Dis. 2007;2(1):1–6.

14. Sticova E, Jirsa M. ABCB4 disease: Many faces of one gene deficiency. Ann Hepatol [Internet]. 2020;19(2):126–33. Available from: 10.1016/j.aohep.2019.09.010

15. Mínguez Rodríguez B, Molera Busoms C, Martorell Sampol L, García Romero R, Colomé Rivero G, Martín de Carpi J. Heterozygous mutations of ATP8B1, ABCB11 and ABCB4 cause mild forms of Progressive Familial Intrahepatic Cholestasis in a pediatric cohort. Gastroenterol Hepatol [Internet]. 2022 Oct;45(8):585–92. Available from: https://linkinghub.elsevier.com/retrieve/pii/S0210570521003290

16. Dixon PH, Williamson C. The pathophysiology of intrahepatic cholestasis of pregnancy. Clin Res Hepatol Gastroenterol [Internet]. 2016;40(2):141–53. Available from: 10.1016/j.clinre.2015.12.008

17. Turunen K, Helander K, Mattila KJ, Sumanen M. Intrahepatic cholestasis of pregnancy is common among patients’ first-degree relatives. Acta Obstet Gynecol Scand. 2013;92(9):1108–10.

18. Turro E, Astle WJ, Megy K, Gräf S, Greene D, Shamardina O, et al. Whole-genome sequencing of patients with rare diseases in a national health system. Nature [Internet]. 2020 Jul 2;583(7814):96–102. Available from: https://www.nature.com/articles/s41586-020-2434-2

19. Bacq Y, Sentilhes L. Intrahepatic cholestasis of pregnancy: Diagnosis and management. Clin Liver Dis. 2014;4(3):58–61.

20. Hagenbeck C, Hamza A, Kehl S, Maul H, Lammert F, Keitel V, et al. Management of Intrahepatic Cholestasis of Pregnancy: Recommendations of the Working Group on Obstetrics and Prenatal Medicine - Section on Maternal Disorders. Geburtshilfe Frauenheilkd. 2021;81(8):922–39.

21. Marschall HU, Katsika D, Rudling M, Einarsson C. The genetic background of gallstone formation: An update. Biochem Biophys Res Commun [Internet]. 2010 May;396(1):58–62. Available from: https://linkinghub.elsevier.com/retrieve/pii/S0006291X10003773

22. Geenes V, Williamson C. Intrahepatic cholestasis of pregnancy. World J Gastroenterol. 2009;15(17):2049–66.

23. Wood AM, Livingston EG, Hughes BL, Kuller JA. CME Review Article. Pediatr Emerg Care. 2018;34(1):59–60.

24. Gudbjartsson DF, Helgason H, Gudjonsson SA, Zink F, Oddson A, Gylfason A, et al. Large-scale whole-genome sequencing of the Icelandic population. Nat Genet. 2015;47(5):435–44.

25. Stättermayer AF, Halilbasic E, Wrba F, Ferenci P, Trauner M. Variants in ABCB4 (MDR3) across the spectrum of cholestatic liver diseases in adults. J Hepatol. 2020;73(3):651–63.

26. Loomes KM, Squires RH, Kelly D, Rajwal S, Soufi N, Lachaux A, et al. Maralixibat for the treatment of PFIC: Long-term, IBAT inhibition in an open-label, Phase 2 study. Hepatol Commun. 2022;6(9):2379–90.

27. Thompson RJ, Arnell H, Artan R, Baumann U, Calvo PL, Czubkowski P, et al. Odevixibat treatment in progressive familial intrahepatic cholestasis: a randomised, placebo-controlled, phase 3 trial. Lancet Gastroenterol Hepatol. 2022;7(9):830–42.

28. Jacquemin E. Progressive familial intrahepatic cholestasis. Clin Res Hepatol Gastroenterol [Internet]. 2012;36(SUPPL.1):S26–35. Available from: 10.1016/S2210-7401(12)70018-9

29. Finer S, Martin HC, Khan A, Hunt KA, Maclaughlin B, Ahmed Z, et al. Cohort Profile: East London Genes & Health (ELGH), a community-based population genomics and health study in British Bangladeshi and British Pakistani people. Int J Epidemiol. 2020;49(1):20–21I.

30. Saarinen K, Färkkilä M, Jula A, Erlund I, Vihervaara T, Lundqvist A, et al. Enhanced liver Fibrosis® test predicts liver-related outcomes in the general population. JHEP Reports [Internet]. 2023;5(7):100765. Available from: 10.1016/j.jhepr.2023.100765

31. Tamaki N, Kurosaki M, Huang DQ, Loomba R. Noninvasive assessment of liver fibrosis and its clinical significance in nonalcoholic fatty liver disease. Hepatol Res. 2022;52(6):497–507.

32. Berzigotti A, Tsochatzis E, Boursier J, Castera L, Cazzagon N, Friedrich-Rust M, et al. EASL Clinical Practice Guidelines on non-invasive tests for evaluation of liver disease severity and pro gnosis – 2021 update. J Hepatol. 2021;75(3):659–89.

33. Friedrich-Rust M, Ong MF, Martens S, Sarrazin C, Bojunga J, Zeuzem S, et al. Performance of Transient Elastography for the Staging of Liver Fibrosis: A Meta-Analysis. Gastroenterology. 2008;134(4):960–74.

34. Sasso M, Tengher-Barna I, Ziol M, Miette V, Fournier C, Sandrin L, et al. Novel controlled attenuation parameter for noninvasive assessment of steatosis using Fibroscan®: Validation in chronic hepatitis C. J Viral Hepat. 2012;19(4):244–53.

35. Boursier J, Zarski JP, de Ledinghen V, Rousselet MC, Sturm N, Lebail B, et al. Determination of reliability criteria for liver stiffness evaluation by transient elastography. Hepatology. 2013;57(3):1182–91.

36. Avena A, Puggelli S, Morris M, Cerny A, Andrade AR, Pareti E, et al. ABCB4 variants in adult patients with cholestatic disease are frequent and underdiagnosed. Dig Liver Dis [Internet]. 2021 Mar;53(3):329–44. Available from: https://linkinghub.elsevier.com/retrieve/pii/S1590865820310860

37. Mareux E, Lapalus M, Ben Saad A, Zelli R, Lakli M, Riahi Y, et al. In Vitro Rescue of the Bile Acid Transport Function of ABCB11 Variants by CFTR Potentiators. Int J Mol Sci. 2022;23(18).

38. Macpherson I, Abeysekera KWM, Harris R, Mansour D, McPherson S, Rowe I, et al. Identification of liver disease: Why and how. Frontline Gastroenterol. 2022;367–73.

39. Verkade HJ, Felzen A, Keitel V, Thompson R, Gonzales E, Strnad P, et al. EASL Clinical Practice Guidelines on genetic cholestatic liver diseases. J Hepatol [Internet]. 2024;81(2):303–25. Available from: 10.1016/j.jhep.2024.04.006

40. Wang HH, Portincasa P, Liu M, Wang DQH. Genetic Analysis of ABCB4 Mutations and Variants Related to the Pathogenesis and Pathophysiology of Low Phospholipid-Associated Cholelithiasis. Genes (Basel). 2022;13(6).

41. van Mil SWC, van der Woerd WL, van der Brugge G, Sturm E, Jansen PLM, Bull LN, et al. Benign recurrent intrahepatic cholestasis type 2 is caused by mutations in ABCB11. Gastroenterology [Internet]. 2004 Aug;127(2):379–84. Available from: https://linkinghub.elsevier.com/retrieve/pii/S0016508504007759

42. Monrose E, Bui A, Rosenbluth E, Dickstein D, Acheampong D, Sigel K, et al. Burden of Future Liver Abnormalities in Patients With Intrahepatic Cholestasis of Pregnancy. Am J Gastroenterol. 2021;116(3):568–75.

43. Sticova E, Jirsa M, Pawłowska J. New Insights in Genetic Cholestasis: From Molecular Mechanisms to Clinical Implications. Can J Gastroenterol Hepatol. 2018;2018.

44. Nurm M, Reigo A, Nõukas M, Leitsalu L, Nikopensius T, Palover M, et al. Do Biobank Recall Studies Matter? Long-Term Follow-Up of Research Participants With Familial Hypercholesterolemia. Front Genet. 2022;13(July):1–11.

45. Luo L, Aubrecht J, Li D, Warner RL, Johnson KJ, Kenny J, et al. Assessment of serum bile acid profiles as biomarkers of liver injury and liver disease in humans. PLoS One. 2018;13(3):1–17.

