## Supplementary Material for "A Genes & Health pilot recall study of intrahepatic cholestasis of pregnancy and cholestatic liver disease"

**Supplementary Table 1. Summary of volunteers with significant loss of function variants invited for recall by genotype**

| **Volunteers (n)** |  | **Loss of function recall** | | | | | | |
| --- | --- | --- | --- | --- | --- | --- | --- | --- |
|  | **Gene** | **Type** | **Variants** | **Effect** | **Zygosity** | **Transcript** | **GnomAD AF** | **G&H AF** |
| 10 | *ABCB4* | LoF | S99x | Frameshift | het | ENSP00000395716.1:p. Ser99LeufsTer11 | 0.00039970 | 0.00336538 |
| 1 | *ABCB4* | LoF | F758X | Frameshift | het | ENSP00000392983.1:p. Leu759TyrfsTer38 | . | 0.00009610 |
| 1 | *ABCB4* | LoF | Lys30GlyfsTer7 | Frameshift | het | ENSP00000392983.1:p. Lys30GlyfsTer7 | 0.00000408 | 0.00020243 |
| 1 | *ABCB4* | LoF | R595* | Stop gained | het | ENSP00000392983.1:p. Arg595Ter | 0.00001627 | 0.00009593 |
| 1 | *ABCB11* | LoF | A1044x | Frameshift | het | ENSP00000497931.1:p. Ala1044LeufsTer53 | . | 0.00009562 |
| 1 | *ABCB11* | LoF | c.2611-2A>T | Splice-acceptor variant | het | ENST00000263817.7:c.2611- 2A>T | 0.00000407 | 0.00057870 |
| 1 | *ABCB11* | LoF | W239x | Stop gained | het | ENSP00000497931.1:p. Trp239Ter | . | 0.00009566 |

**Abbreviations:** AF – Allele frequency, G&H – Genes and Health, GnomAD- Genome aggregation database, Het-Heterozygous, LoF- Loss of function. Inclusion criteria (< 1% MAF).

**Supplementary Table 2. Summary of volunteers with variants invited for recall by phenotype**

| **Volunteers (n)** | **Phenotype recall** | | | | | | | | |
| --- | --- | --- | --- | --- | --- | --- | --- | --- | --- |
|  | **Phenotype** | **Gene** | **Type** | **Variants (SNV)** | **Zygosity** | **Transcript** | **dbSNP** | **GnomAD AF** | **G&H AF** |
| 1 | ICP | *ABCB4* | SNV | G1254S | het | ENSP00000496956.1:p. Gly1254Ser | rs781315185 | 0.00003656 | 0.00028843 |
| 1 | ICP | *ABCB4* | SNV | P1050S # | het | ENSP00000497931.1:p. Asp1284Asn |  |  | 0.00019146 |
| 1 | ICP | *ABCB4* | SNV | A833T # | het | ENSP00000496956.1:p. Ala833Thr |  |  | 0.00009638 |
| 1 | ICP | *ABCB4* | SNV | N510S | het | ENSP00000496956.1:p. Asn510Ser | rs375315619 | 0.00019110 | 0.00057394 |
| 1 | ICP | *ABCB4* | LOF | S99x | het | ENSP00000395716.1:p. Ser99LeufsTer11 |  | 0.00039970 | 0.00336538 |
| 1 | ICP | *ABCB11* | SNV | D1284N # | het | ENSP00000497931.1:p. Asp1284Asn | rs766784155 | 0.00001228 | 0.00028780 |
| 1 | ICP | *ABCB11* | SNV | R1050H | het | ENSP00000497931.1:p. Arg1050His | rs72549398 | 0.00000421 | 0.00019135 |
| 1 | ICP | *ABCB11* | SNV | V284A | het | ENSP00000497931.1:p. Val284Ala | rs200739891 | 0.00026040 | 0.00009558 |

**Abbreviations:** AF – Allele frequency, dbSNP - Single nucleotide polymorphism database, G&H – Genes and Health, GnomAD - Genome aggregation database, Het - Heterozygous, ICP – Intrahepatic cholestasis of pregnancy, MAF- Minor allele frequency, LoF - Loss of function, SNV- Single nucleotide variant. Inclusion criteria (< 1% MAF).
